# Evaluating the effectiveness of vestibular and ocular motor function assessments in detecting driver sleepiness: A Protocol Paper

**DOI:** 10.64898/2026.02.18.26346511

**Authors:** Alisha Guyett, Claire Dunbar, Nicole Lovato, Katrina Nguyen, Kelsey Bickley, Duc Phuc Nguyen, Amy Reynolds, Maslin Hughes, Hannah Scott, Robert Adams, Leon Lack, Peter Catcheside, Lucia Pinilla, Jennifer Cori, Mark E. Howard, Clare Anderson, David Stevens, Darah-Bree Bensen-Boakes, Ashley Montero, Nicole Stuart, Andrew Vakulin

## Abstract

**Background:** Prolonged wakefulness, restricted sleep, and circadian factors can impact driving performance and road safety. Currently, there are no effective objective roadside tests to detect the state of driver’s sleepiness during or prior to driving, or predict future driving impairment risk. This paper reports on an extended wakefulness protocol used to determine if a portable virtual reality device to administer vestibular-ocular motor function (VOM) tests can effectively detect 1) driver’s state of sleepiness during or just prior to driving, and 2) predict trait sleepiness and future driving risk.

**Methods:** Fifty healthy adults with regular sleep within 9pm to 8am were recruited for an experimental laboratory procedure which involved two phases: an initial overnight sleep study, and a subsequent period of extended wakefulness lasting ~29 hours. During the wakefulness phase, participants undertook neurobehavioural testing, a simulated driving test, and repeat assessments of VOM to establish if ocular markers can predict sleepiness state and sleepiness-related performance impairments (Trial registry ACTRN12621001610820).

**Discussion:** This protocol outlined a study that aimed to establish the sensitivity of VOM test the effects of extended wakefulness and circadian phase on driver state and trait sleepiness and subsequent sleepiness-related driving impairment. Furthermore, the protocol aims to define the best VOM predictors to identify driver sleepiness state (road side testing and pre-drive assessments) and sleepiness trait (predicting future driving risk) to establish proof of concept for its potential application as a roadside, pre-drive and general sleepiness related fitness to drive test.

## Introduction

Excessive sleepiness (i.e., the inability to maintain wakefulness or alertness) increases the risk of a motor vehicle accident by 250% (Bioulac et al., 2017). Driver sleepiness is implicated in up to 20% of all motor vehicle crashes, increasing to 30% for single vehicle crashes (de Mello et al., 2008). Given the importance of alertness for driving performance and road safety, roadside or workplace tests for detecting driver sleepiness are a potential high-value target and valuable tool for promoting road safety. Unlike roadside or workplace tests for detecting impairments from drugs and alcohol (Boorman & Papafotiou, 2007), which aim to enhance public safety by identifying intoxicated individuals before they cause accidents, validated approaches to objectively evaluate sleepiness-related driving impairments are lacking. Blood alcohol assessments target driving impairment by assessing blood alcohol levels associated with unacceptable impairment risk. In an analogous manner, this study sought to test the potential utility of vestibular-ocular motor (VOM) function test as a practical objective test that may be sensitive to sleepiness, and sleepiness-related driving impairment. Sleep related driving accidents can arise from sleepiness related inattention and neurocognitive deficits that reduce task performance with increased error rates, and complete failure to sustain wake (De Mello et al., 2013). Such microsleep events typically result in the absence of last minute evasive manoeuvres (braking/ swerving) to increase the likelihood of head on or other high speed collisions with higher fatality risks than alcohol or drug-related accidents (Thomas et al., 2021). Thus, practical, and reliable methods to evaluate and detect the risk of neurobehavioural impairment from sleepiness are highly desirable.

Sleep loss has a widespread detrimental impact on performance including reduced vigilance to external stimuli, instability in sustaining attention, and inability to inhibit distraction (Al-Mekhlafi et al., 2020; Lim & Dinges, 2008). Extrapolating these findings to driving a road vehicle is problematic but supports that sleepiness may promote errors such as failure to stop at a red light, and slowing of reactions to external events (e.g., brake lights, pedestrians, variation in speed and drifting out of lane).

Sleepiness associated with driving impairments can be detected via physiological, behavioural, and cognitive tasks. Self-reported sleepiness via single-item scales and questionnaires are commonly used in experimental settings (Anderson et al., 2023; Cai et al., 2021; Cai et al., 2023). However, there is considerable potential for bias due to the self-report nature of these assessments, particularly when there are perceived (and/or actual) consequences associated with reporting sleepiness. Evidence also suggests that individuals are unable to accurately perceive objective impairment related to their level of sleepiness when sleep deprived (McCauley et al., 2021). Furthermore, self-reported sleepiness may not be sufficient to influence a driver’s decisions to stop driving or apply alternative safety countermeasures to promote wake, or to simply continue driving with sleepiness itself potentially impairing rational decision making around risks (Watling et al., 2015). Given the major limitations of self-report evaluations, objective sleepiness assessments are more desirable for evaluating sleepiness-related driving impairments and risks.

Objective measurements of sleepiness-related driving impairments can be obtained through either direct measurement of driving performance itself via vehicular measurements (e.g., steering deviation/lane departures) or more specifically target the driver’s behaviour and sensorimotor functions (e.g., camera-based assessment of head and ocular movements). A meta-analysis of 15 papers utilising lane tracking (Soares et al., 2020) to evaluate the effects of drowsiness on driving performance found that sleepy individuals showed greater variability in lane position and a higher incidence of lane departures compared to alert drivers. However, this approach requires the vehicle to be fitted with costly and sophisticated vehicle tracking technology with limited data to support measurement reliability and utility. Eye-tracking via electrooculography (EOG) can be used to measure specific patterns of vertical and horizontal directional changes in the pupil position (i.e., eye rolling) and eyelid closures (Hu & Lodewijks, 2021), which are associated with driving impairment and crashes in laboratory and instrumented on-road vehicle studies (Cori et al., 2023; Jackson et al., 2016).

Electroencephalography and electrocardiography have also been used to record sleepiness-related changes in brain and heart activity (Balandong et al., 2018). These require continuous recording via electrodes, which is somewhat intrusive and impractical for regular use and may impact driving performance via discomfort. Cardiac features such as heart rate may also be too non-specific to be useful for evaluating sleepiness. Non-contact devices are favourable as they minimise interference with the driver (Liu et al., 2022), such as in car infra-red cameras to detect sleepiness-related behaviours such as yawning, head posture (slumping of the head), and slow eye rolling. The inherent need to capture facial features through camera recording has raised privacy concerns, which may prevent users from engaging with this technology. Therefore, these existing markers of driver sleepiness may be too impractical for use in most real-world contexts (Anderson & Horne, 2006; Lerman et al., 2017).

Vestibular-ocular motor (VOM) function assessments are ocular-based tools that may be more practical to detect objective sleepiness (Anderson et al., 2013). VOR (Vestibular Ocular Reflex) assessments include smooth pursuit (eye tracking ability), markers of the ability to compensate for head movement and maintain focus, saccades (rapid eye movements between visual fixations), and nystagmus (repetitive and uncontrolled eye movements). Several of these ocular measures appear to be sensitive to sleepiness, including percentage or duration of eye closures (Åkerstedt et al., 2005; Hu & Lodewijks, 2021) and blink durations (Cori et al., 2019; Stern et al., 1994). VOM and near point convergence scores also appear to be sensitive to fatigue (Ratka et al., 2020). Ratka et al., (2020) showed that VOM scores were sensitive to exertional fatigue, and concluded that clinicians using VOM for clinical practice should allow rest prior to assessment to allow recovery (Ratka et al., 2020). Until recently, VOMS assessments required specialised facilities to administer (Brandt & Strupp, 2005). With rapid growth in virtual reality technology, VOM can now be conducted with portable equipment readily deployable to field settings. Thus, the purpose of this study protocol was to evaluate the sensitivity of VOM based measurements to sleepiness and sleepiness-related driving simulator task performance decrements induced by extended wakefulness.

### Project aims

This study protocol was designed to determine the sensitivity of VOM test metrics to driver state and trait sleepiness and consequent driving simulator impairment during extended wakefulness with the aim of defining the best VOM predictors of driver sleepiness and driving impairment. Specifically, the primary study aims are to:

1. Define the top VOM predictors of driver sleepiness state and driving impairment using VOM assessments occurring straight after each driving assessment throughout the extended wakefulness. This simulates a road-side test scenario where a driver is pulled over after a period of driving and their sleepiness state tested
2. Define the top VOM predictors of driver sleepiness state and subsequent driving impairment using VOM test occurring just before initiation of driving. This simulates a fitness to drive assessment scenario where driver’s sleepiness state is tested before driving begins and predicts the subsequent driving impairment.

A secondary aim is to determine if VOM assessment at baseline when drivers are rested (<16hrs of wakefulness) can predict and discriminate between drivers who subsequently exhibit vulnerability versus resistance to driving and alertness failure following (20+hrs) extended wakefulness. This simulates a scenario where a driver requires a generalised fitness to drive assessments as part of occupational or clinical fitness to drive assessments.

Whilst multiple vestibular ocular measures were assessed for exploratory purposes, based on previous research it was anticipated that the VOM metrics with the highest likelihood to predict neurobehavioral impairment will be higher anti-saccade error rates (mistakes made), increased saccadic velocity (Bocca et al., 2014; Fransson et al., 2008; Hellmuth et al., 2012; Zils et al., 2005) and reduced smooth pursuit velocity gain (Fransson et al., 2008; Meyhöfer et al., 2017; Tong et al., 2014) and these will predict higher rates of driving simulator impairment. Based on previous research VOM metrics extracted from the saccadic and anti-saccadic tasks were of particular interest but all VOM metrics were considered as potential predictors of state and trait sleepiness and driving impairment. To strengthen the validity of the top VOM predictors of state and trait sleepiness and driving impairment we also examined them in relation to other established non driving markers of alertness (psychomotor vigilance test, PVT, Karolinska drowsiness test, KDT, Karolinska Sleepiness Scale, KSS, and electroencephalographic, EEG, and core and skin temperature changes as circadian markers) to demonstrate that our findings do not relate to a particular driving simulator but are representative of general sleepiness as a construct.

Several exploratory aims of the study encompassed elements of the protocol unrelated to the primary or secondary aims including electroencephalographic assessment of the sleeping brain, executive function performance, circadian rhythm tracking and subjective sleepiness are not described in detail in the current paper for succinctness.

### Hypotheses

The protocol was primarily designed in an exploratory fashion to test the hypotheses that:

1. Ocular markers of volitional and reflex eye-movements (anti-saccades, saccades, smooth pursuit) will be among the top predictors of state and trait sleepiness and driving performance decrements (increased lane deviation and crashes during simulated driving), attentional lapses and reduced reaction time (PVT) during extended wakefulness.
2. The baseline VOM metrics assessed during rested state prior to extended wakefulness (<16hrs awake) will be predictive of future subsequent sleepiness, driving and psychomotor impairments following 20+hrs of extended wakefulness.

## Methods

### 2.1 Study Design

This manuscript describes the study protocol for an extended wakefulness in-laboratory study designed to examine the relationships between vestibular ocular measures, state sleepiness and simulated driving performance. The trial was registered prospectively with the Australian and New Zealand Clinical Trails Registry (ACTRN12621001610820, 26 November 2021). The study was approved by the Flinders University’s Human Research Ethics Committee (project number 4648, approved on the 15.09.2021), and all participants provided informed written consent to participate in this study and for their de-identified data to be reported in publications. Data collection was completed from 2021 to 2023.

### 2.2 Facilities

The experimental procedures were conducted at the Nick Antic Sleep Laboratory (Flinders Health and Medical Research Institute: Sleep Health (FHMRI: Sleep Health / Adelaide Institute for Sleep Health). This facility allows for tight control of light (off, <1lux during sleep opportunities, and dim warm light <10lux throughout wake) and temperature (22°C), including in six sound-attenuated bedrooms, each equipped with a king-single bed and ensuite bathroom, and a shared lounge area for participant use during waking hours.

### 2.3 Study Participants

Participants were recruited from individuals registered on the FHMRI: Sleep Health laboratory volunteer registry who self-identified as healthy sleepers and met preliminary eligibility criteria, and via online and community advertisements and through word of mouth. Individuals who expressed interest in study participation, and met initial eligibility criteria, completed further online screening before being sent the participant information sheet and providing written consent to participate if all eligibility criteria were met.

### 2.4 Exclusion and Inclusion Criteria

Basic screening and demographic questions were asked in a general screening questionnaire to assess eligibility including age, weight, height, date of birth, work status, driving license status, habitual sleep timing, current health diagnoses including psychiatric and sleep disorders, health status, current medication use and/or illicit substance use, average alcohol and caffeine intake.

#### 2.2.1 Inclusion

Participants met inclusion criteria if they were aged ≥18 years to 75 years, able to provide written informed consent, had a full current car license, reported driving at least 3 hours per week, reported a regular sleep schedule; bedtime between 20:00-00:00; wake time between 05:00-09:00, reported a typical sleep duration ranging between 6-10 hours; were fluent in both reading and speaking English, recorded an insomnia severity Index score (ISI) <10 indicating minimal insomnia symptoms, recorded and Epworth Sleepiness Scale score (ESS) <10 indicating normal daytime sleepiness, recorded an Obstructive Sleep Apnoea (OSA) 50 screening questionnaire < 5 indicating low likelihood of sleep apnoea. During the study recruitment inclusion for older adults >35yrs to 75yrs was revised in line with expected normal differences for sleep in relation to the older adults. A case by case inclusion criteria was adopted for individuals who scored above the recommended cut offs for ISI, ESS and OSA, as sleep disorders and difficulties are more common in adults aged >35 years (45% population (McArdle et al., 2022)), and to take into account the OSA-50 inflating age criteria. Participants were still required to fall <14 on the ISI, <16 on the ESS (ie., less than severe excessive daytime sleepiness).

#### 2.2.2 Screening Questionnaires

Insomnia symptoms and impacts on daily functioning were assessed via the insomnia severity index (ISI), a 7-item self-report questionnaire (Bastien et al., 2001). Each item is rated on a Likert-type scale ranging from 0 to 4 (higher scores indicate greater severity of insomnia symptoms) for a total score between 0 and 28. A total score of <10 was considered as not presenting clinical symptoms of insomnia and therefore used as a screening cut off measure for recruitment (Morin et al., 2011).

Sleepiness was evaluated using the Epworth sleepiness scale (ESS) an 8-item, self-report questionnaire assessing an individual’s propensity to fall asleep in various ‘daily life’ situations (Johns, 1991) (e.g., sitting and reading, watching television, or being a passenger in a vehicle). Participants rate their likelihood of dozing off in each scenario on a scale from 0 (no chance of dozing) to 3 (high chance of dozing), with a total score ranging from 0 to 24 (higher scores indicating greater daytime sleepiness). A total score of <10 is considered as not presenting with excessive daytime sleepiness.

The four-item OSA-50 screening questionnaire was used to help screen out OSA (Chai-Coetzer et al., 2011) on the basis of self-reported snoring (a score of 3 if positive), cessation of breathing during sleep (a score of 2 if positive), and age (a score of 2 if ≥ 50 years), and a measurement of obesity (a score of 3 if waist circumference is > 102 cm for males and > 88 cm for females). Individuals scoring ≥ 5 out of 10 were considered high risk of OSA and excluded from the current study which aimed to recruit healthy sleepers.

#### 2.2.3 Exclusion

Participants were excluded from the study if they reported current physical or mental health conditions, including diagnosed sleep conditions, that may alter sleep; a BMI >35kg/m^2^ indicating heightened risk of sleep disordered breathing associated with weight, a History of traumatic brain injury, stroke, or neurodegenerative disorders; active use of medication or illicit substances that may alter sleep; elevated alcohol and caffeine use (>4 times alcoholic and/or caffeine drinks per day; OR >10 alcoholic drinks per week) with the potential to alter sleep; smoking dependence (non-casual smoking) which would prevent them from being able to refrain from smoking for the duration of the study; pregnancy, lactating, or caring for a newborn (12 months and under) which may alter sleep unrelated to the study; recent overnight shift workers who may have an already accumulated sleep debt; recent time-zone travel encompassing >2 zones (in the previous 2 months) which may have altered normal circadian alignment.

### 2.5 Measure/Materials

All scales and performance tests used in this study were administered in English, via online survey delivery program Qualtrics.

#### 2.5.1 Sleepiness and Performance Measures

Participants attended the laboratory in the evening for direct polysomnographic (PSG) evaluation of sleep on the first night of the study protocol. PSG was used to directly evaluate sleep on the first night to assess sleep quality and screen for undiagnosed sleep disorders such as obstructive sleep apnoea (OSA) (Caton, 1970). PSG is widely recognised as the gold standard for diagnosing sleep-related breathing disorders (Rundo & Downey, 2019) and is widely used in laboratory sleep research. Electroencephalography (EEG) was acquired using the Grael 4K and ProFusion 4 EEG software from Compumedics Ltd., (Abbotsford, Vic). Frontal, central, and occipital EEG activity were recorded through gold-cup electrodes placed on specific scalp locations using the ten20 system (F3/Fz/F4/C3/Cz/CPz/Pz/C4/O1/Oz/O2) referenced to mastoids (M1 and M2).

Chin electromyography (EMG) was measured to assess skeletal muscle activity, while left and right EOG was employed to measure eye movements. EEG electrode impedances was required to be below 5 kOhms to be considered sufficiently clean for analysis. A single trained sleep technologist, unaware of participant identities, scored the sleep studies in 30-second epochs into rapid eye movement (REM) sleep and non-REM sleep stages based on the American Academy of Sleep Medicine scoring criteria (Iber et al., 2007).

In addition to PSG, participants were equipped with additional sensors to measure breathing during sleep. These sensors include bands around the chest (thoracic) and stomach (abdominal) to monitor expansion and contraction of the lungs, as well as a nasal cannula and thermistor to measure and record nasal airflow. Sensors were also placed on the sides of each calf muscle to track major muscle movement throughout the night, and an oximeter was placed on the non-dominant index finger to measure arterial oxygen saturation and finger pulse oximetry. Electrocardiography (ECG) electrodes were applied to the chest to measure heart rate continuously throughout the protocol. After the overnight sleep, all non-essential PSG components, apart from monitoring brain waves (via EEG), chin electromyography (EMG), heart rate (via ECG) and eye movements (via EOG) during wakefulness, was removed.

#### 2.5.2 Vestibular-Ocular Motor (VOM) test

The Neuroflex VOM device uses virtual reality goggles to record both head and eye motion using infra-red eye-tracking incorporated into a virtual reality headset to specifically target VOR related measures (Figure 1) (Anderson et al., 2013; Mucha et al., 2014). This device measures multiple vestibular/ocular behaviours in under 10 minutes and has recently been used to objectively assess neurological function after a traumatic brain injury (Stevens et al., 2023) and in the assessment of reading and grammatical decisions (Mirault et al., 2020) using movement-based biometrics (Lohr et al., 2018). Each test evaluates specific aspects of eye movement, coordination, and reflexes, providing insights into the functioning of the visual, vestibular, and cognitive systems. Deviations from expected measurements can indicate weaknesses or disorders in these systems. The VO test battery includes eight different test protocols. The Supplementary File contains a list of VOMS variables obtained from the assessments described below.

1. Smooth Pursuit (Head Free): This test involved following a slow-moving target with both eyes and head, promoting coordinated eye-head movements.
2. The target moves in two dimensions, and the subject occasionally makes corrective jerks to maintain focus. Healthy subjects should exhibit minimal vergence (eye deviation), ensuring clear vision.
3. Smooth Pursuit (Head Fixed): Similar to the previous test, but the head is fixed. The subject must track a moving target at a fixed distance with coordinated eye movements. This test evaluates visual and vestibular system coordination.
4. Active Visual VOR (Horizontal and Vertical): This evaluated the vestibulovisual reflex by combining visual, vestibular, and neck proprioception inputs. The subject tracks a moving target while coordinating eye movements with head and neck proprioception. Deficits may indicate weaknesses in the reflex system.
5. Saccades: This assesses rapid eye movements (saccades) to new targets without head movement. It analyses aspects such as reaction time, precision, and vergence. A minimal vergence angle is expected for clear vision during saccades.
6. Anti-Saccades: In this cognitive test, the subject must look away from a briefly presented target, testing the visual cortex’s function. Reaction time and deviations are measured, and excessive deviations can lead to blurry vision.
7. OKN Nystagmus: The Optokinetic Nystagmus test involved tracking scrolling visual stimuli to trigger involuntary eye movements. Results are analysed similarly to RVVOA (active visual VOR). Healthy peripheral retina vision results in similar measurements.
8. Spontaneous Nystagmus: This test detected involuntary eye movements without external stimuli. Imbalances in gaze orientation, potentially from trauma or neurological issues, can cause nystagmus. The test measured the frequency of jerks, eye drift, and tremor.

**Figure 1.**
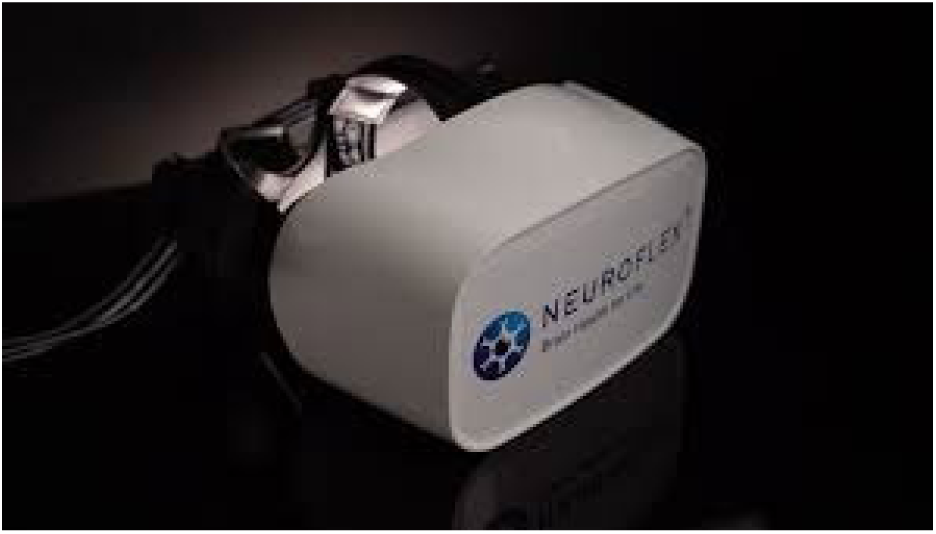
An example of the Neuroflex© virtual reality device employing the FOVE goggles with infrared eye detection.

See supplementary file for additional information.

#### 2.5.3 The AusEd Driving Simulator

Driving performance was assessed using the AusEd driving simulator (Woolcock Institute for Medical Research, Sydney, Australia)(Desai et al., 2007). The AusEd driving simulation is a computer-based driving task designed to represent driving on a dual-lane highway at night-time, whilst using low-beam lights and staying within a speed limit of 60-80kph. Drive duration was set to 60 minutes, with 5 trucks appearing during the drive approximately every ten minutes. At these pre-determined time points, slow-moving trucks were presented ahead of the driver in which case they are asked to brake (registered at 90% of full pedal depression) their vehicle until the truck disappeared to measure reaction time (time taken for individual to brake in response to truck appearing). There are no other vehicles or traffic markers present other than evenly placed reflective markers on the edges and in the middle of the road. The layout of the road and time of presentation of trucks were identical for each drive. An approximately 60 dB low-frequency engine noise was played for the length of the drive(Desai et al., 2006).

The primary AusEd outputs were as follows: Steering deviations from the centre of the left hand lane, steering deviations from the median lane position during the drive (allows for differences in the interpretation of “stay in the centre of the lane position”), speed deviations from the required 60-80kph speed, represents a divided attention task due to the speedometer being beyond the sight of the road, response reaction time to the truck presentation in which participants are asked to brake, number of instances in which the participant crashes the vehicle. The AusEd is sensitive to performance impairment from factors such as sleep deprivation (Desai et al., 2007). (Desai et al., 2007). While the AusEd simulator was the most appropriate choice for the current study we recognise the need for future on road testing for external validity.

#### 2.5.4 Psychomotor Vigilance Task (PVT)

The PVT task was used as a measure of sustained attention (Wilkinson & Houghton, 1982). A computer-based 10-minute version of the task, downloaded from the Inquisit Millisecond Test Library was administered to participants. Participants were instructed to press the space bar as quickly as possible when a target stimulus (a millisecond time counter) appears on the computer screen. Participants were informed that speed and accuracy are equally important on all tasks. The inter-stimulus interval varied randomly between 2-10s. Lapses were defined as instances of ≥500ms reaction time. The PVT has been widely employed in circadian rhythm, sleep restriction, and extended wake protocols and is sensitive to total sleep deprivation, chronic and/or partial sleep deprivation, and circadian disruption (Van Dongen & Dinges, 2005). The PVT test is an established sensitive and validated essay of sleep loss(Basner & Dinges, 2011; Basner et al., 2011) and has high test-retest reliability, with intraclass coefficients above 0.8 for both lapses and median response times (Dorrian et al., 2004).

#### 2.5.5 Karolinska drowsiness test (KDT)

The KDT provided an indication of the individual’s level of sleepiness through a standard procedure enabling assessment of sleepiness through brain (EEG) and ocular (EOG) signals. The test consisted of three x 2.5-minute phases: with eyes open, with eyes closed, and with eyes open again. Participants are instructed to sit quietly throughout and refrain from movement. During the eyes open phase, participants were asked to focus on a large black dot on the wall. EEG signals were recorded during the test, and the analysis of the signals allowed for an assessment of participants’ physiological sleepiness levels (Åkerstedt & Gillberg, 1990). The KDT is associated with sleep loss and circadian disruption(Glos et al., 2014). The KDT is also associated with self-report assessments of sleepiness from the Karolinska Sleepiness Scale (Putilov & Donskaya, 2013).

#### 2.5.6 Withings Sleep Analyser

The Withings Sleep Analyser was used to measure sleep patterns and quality throughout the home-based baseline assessment period of the study (Edouard et al., 2021). The non-intrusive device is placed under the mattress, in line with the upper chest position. Using ballistography via the in-built air bladder, the device estimates movement, heart rate and respiration signals to derive various sleep parameters, including sleep duration, sleep onset and offset times, snoring, and breathing disturbances during sleep. Data is transmitted via Bluetooth to a companion app, providing a detailed report on participant’s sleep quality and patterns. The WSA demonstrates relatively high accuracy for a consumer sleep monitor (Lechat et al., 2022; Scott et al., 2023), and reasonable precision and had high sensitivity (88.0%) and specificity (88.6%) for detecting sleep apnoea(Edouard et al., 2021).

#### 2.5.7 GENEActiv

The GENEActiv device is a wrist-worn, tri-axial accelerometer designed for the objective measurement of physical activity and sleep (Rosenberger et al., 2016). The device is compact, lightweight, and was constructed from durable materials, including a polycarbonate case and a silicone wristband. The accelerometer consists of a MEMS (micro-electro-mechanical system) sensor, which detects acceleration and movement, and a microcontroller unit which records data. The device has a data storage capacity of up to 4GB and was able to record data at a sampling rate of up to 100Hz. The GENEActiv includes a range of features, including a light sensor, a button for user input, and wireless connectivity, which enabled data to be downloaded and analysed remotely. Participants were provided with a GENEActiv device at baseline consent meeting and wore the device for the entire study protocol.

### 2.6 Computer Specifications for Computer Based Performance Measures

The performance tasks were conducted on high-performance gaming computers to ensure minimal latency for the participants. The Dell OptiPlex 7080 was used as the computer system, equipped with 16GB RAM, Intel(R) Core i7-10700 CPU, and NVIDIA GeForce GTX 1660 SUPER 6GB graphics card. These computers have dual monitors (Dell 2319H, 5ms response time), an Alienware 310K mechanical gaming keyboard with 100Hz polling rate (1ms response time), and a Dell Laser Wired Mouse - MS3220. The use of this equipment was critical in reducing the impact of systematic (such as the keyboard being generally slow) and random (such as the computer randomly glitching) biases on the performance data, especially for time-sensitive tasks like the PVT.

### 2.7 Laboratory Questionnaires

#### 2.7.1 Sleep and Health Measures

A comprehensive survey was conducted upon arrival at the participants’ laboratory stay, which collected demographic data, trait sleep measures, and generic motor vehicle data. This survey includes date of birth, biological sex and gender, as well as health data related to handedness and eyesight. Sleep questionnaires included the Pittsburgh Sleep Quality Index (Carpenter & Andrykowski, 1998), the Good Sleeper Scale (Manners et al., 2023), the Morningness Eveningness Questionnaire (Horne & Ostberg, 1976), the Sheehan Disability Scale (Leon et al., 1997), the Short-Form Health Survey (Montazeri et al., 2005), the 21 item Depression, Anxiety and Stress Scale (Lovibond & Lovibond, 1995), and the Fatigue Severity Scale (Valko et al., 2008). In addition to these validated questionnaires, the survey also queried history of workplace and motor vehicle accidents and assessed any potential relation to sleep-impaired performance in the past, allowing the researchers to determine prior likelihood based on previous accidents in these environments.

#### 2.7.2 Sleep Diary

To assess subjective sleep and habitual sleep timing, an online version (via Qualtrics software) of the Consensus Sleep Diary were completed by participants each morning during baseline. Participants self-report questions pertaining to time in/out of bed and minutes awake/asleep in bed per night, enabling the calculation of time in bed, sleep latency, number and duration of awakenings, wake up time and total sleep time (Carney et al., 2012). The Consensus Sleep Diary is well-validated and shows high agreement compared to PSG (kappa = 0.87) and high sensitivity (92.3%) and specificity (95.6%) (Rogers et al., 1993).

#### 2.7.3 Karolinska Sleepiness Scale (KSS)

The KSS is a validated measure of subjective sleepiness and obtains ratings of arousal ranging from subjective alertness to sleepiness (Åkerstedt & Gillberg, 1990). The single-item KSS asks participants to self-report their current sleepiness level on a scale ranging from 1 = extremely alert, to 9 = extremely sleepy. Higher scores on the KSS resemble higher sleepiness/lower alertness. The KSS is sensitive to changes in alertness and sleepiness across various durations of extended wakefulness (Gillberg et al., 1994; Lo et al., 2012).

### 2.8 Participant Controlled Protocol Activities

During the study, food intake was closely monitored and strictly controlled to ensure consistency across all participants. Participants were prohibited from bringing any food or beverages into the laboratory. Given that food consumption increases metabolic rate and can affect core body temperature readings, small meals (~1255kJ per meal) provided to participants at 2-hour intervals. This approach aimed to minimise inaccurate temperature readings that are not associated with circadian rhythms (Monnard et al., 2017). Additionally, the rigorous protocol allowed for modelling techniques to eliminate time periods where artifacts due to food consumption were likely to occur. During the extended wakefulness period temperature and light exposure was kept consistent at 23 degrees and ~5lux to simulate low light exposure experienced on night shifts. Participants were asked to refrain from high intensity exercise but were able to move about the laboratory as needed and were asked to stay mostly in an upright or seated position for most of the protocol.

### 2.9 Protocol

The Figure 2 depicts that Vestibular Ocular Motor assessments were conducted before and after each driving simulation task to predict driving performance (Figure 2A). Specifically, VOM assessments were performed using a virtual reality headset (Figure 2B). The raw data from these assessments were automatically uploaded to the NeuroFlex cloud platform, where 61 VOM metrics across six different domains were computed (Figure 2C). Driving simulator performance was evaluated using two metrics: the steering deviation from the median lane position and the number of crashes. Participants were then classified into two groups—resilient or vulnerable to extended wakefulness—based on these driving performance metrics using cluster analysis (Figure 2D). In the machine learning model (Figure 2E), the 61 VOM metrics served as predictors, while participant groups (resilient vs. vulnerable) were treated as outcome variables. The model was validated using a leave-one-out cross-validation approach (Meijer & Goeman, 2013).

**Figure 2:**
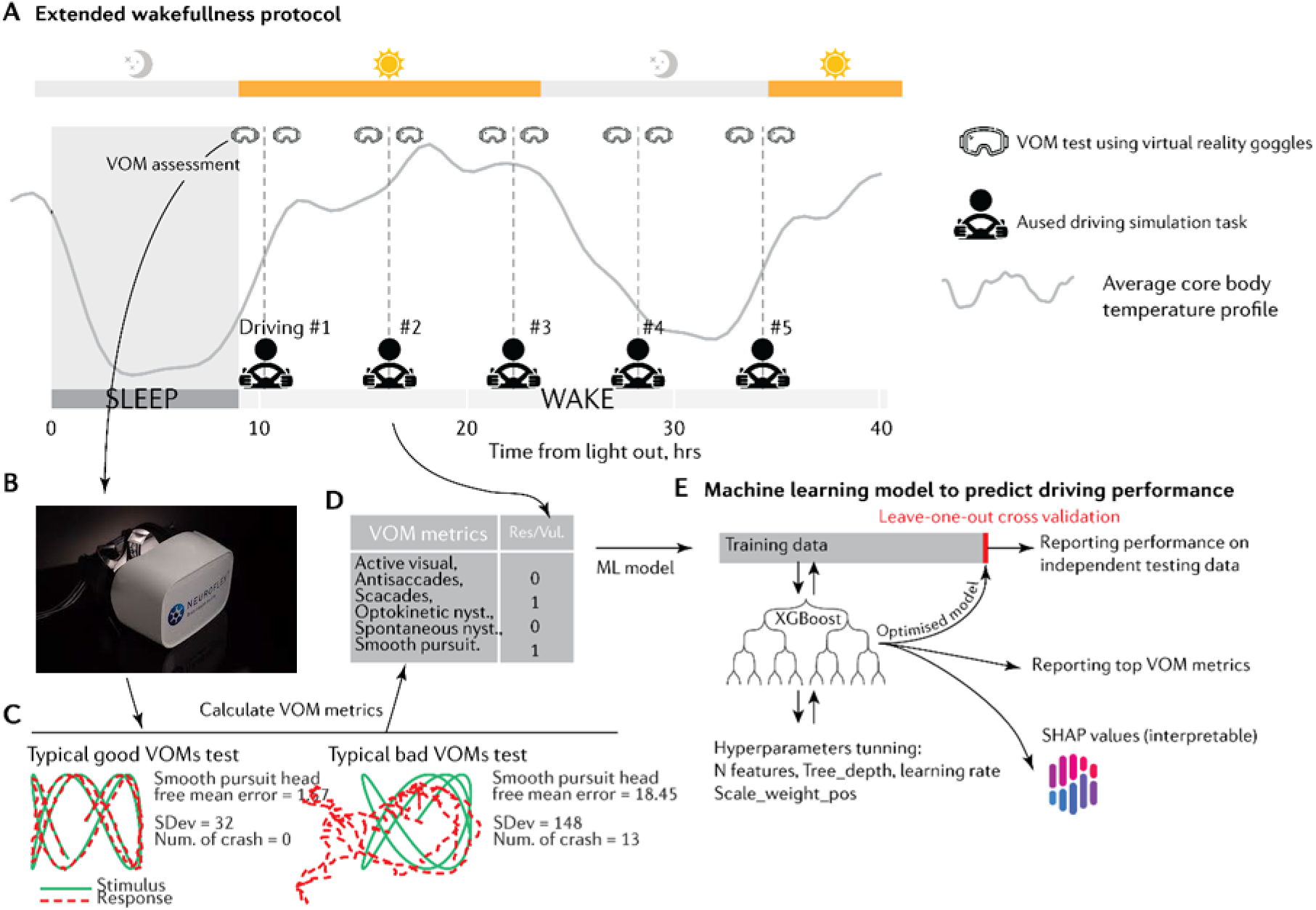
provides an overview of the study protocol and analysis approach/framework. (A) 29hr extended wakefulness protocol during which each participant completed 1hr driving simulator assessments and other vigilance tests at 5 time points throughout extended wakefulness. (B) VOM assessments were conducted pre and post each driving test using virtual reality goggles (NeuroFlex® device). (C) Shows a representative example of the effect of extended wakefulness on one of the VOM domains (smooth pursuit) and the concurrent impact on driving simulator performance. (D) Driving performance was evaluated using two main driving simulator variables: the steering deviation (standard deviation from median driving lane position) and the number of driving crashes. These driving variables from each of the 5 driving tests were used to classify all participants into two groups of drivers who were either vulnerable (impaired) or resilient (unimpaired) to driving impairment. (E) The 61 VOM metrics were then input into an XGBoost (eXtreme Gradient Boosting)(Chen & Guestrin, 2016) machine learning model which was trained using leave-one-out cross validation approach to predict which participants were resilient vs vulnerable to driving impairment. The prediction models were optimized using a Bayesian optimization approach (Diederik, 2014) and Shapley values (SHAP) (Lundberg et al., 2020), were used to rank VOM metric importance (higher absolute Shapley values indicate greater importance of a VOM metric in predicting vulnerable vs resistant drivers).

#### 2.9.1 Initial Laboratory Visit

Participants were given the opportunity to complete all of the laboratory assessments in their full form, with the exception of the driving simulator, which was a 30-minute practice session. The complete set of laboratory assessments took approximately two hours to complete.

#### 2.9.2 Two-week home baseline

During the first week of the baseline period, participants were asked to follow their regular sleep schedule and habits. In the second week of the baseline period, participants were requested to avoid shift work and sleep deprivation in the lead up to the study. During the second week of baseline monitoring, participants were also instructed to avoid caffeinated substances to reduce the likelihood of experiencing withdrawal symptoms during their stay, as stimulants were not permitted in the laboratory. Throughout the entire baseline period, participants’ sleep was monitored using online sleep diaries, which they completed immediately upon waking to note duration, quality and any sleep-related events or disturbances.

#### 2.9.3 Experimental Laboratory Stay

Upon their arrival at the sleep laboratory, participants spent approximately ~39 hours, which included two nights and one and a half days in the laboratory. From 19:00-07:00 on the first night, participants arrived at the laboratory, completed surveys, and were set up with PSG equipment to monitor their sleep. Measurements of core body and skin temperature were also taken as aimed beyond the scope of the primary analysis and further details can be found in the supplementary file for these measures. Participants began their time in bed at the same time as they have been going to bed at home (e.g., 22:00) to maintain consistency with their usual sleep pattern, but had to fall asleep between 21:00 to 00:00. Participants had a 9hr sleep opportunity which determined their wake time the next morning (e.g., 07:00 depicted in Figure 3 as an example)).Participants then began a controlled constant routine protocol period of extended wakefulness that lasts for approximately 29 hours. During this period, participants completed five identical assessment sessions spaced six hours apart (see Figure 3). Each session included a comprehensive performance battery consisting of both major and minor components, ensuring consistent measurement of cognitive and motor function across the sleep deprivation period. This systematic repeated-measures design enables evaluation of all outcomes at baseline (non-sleep deprived) and sleep-deprived states. The complete task battery timing and detailed protocol are provided in supplementary files 3 and 4.

**Figure 3.**
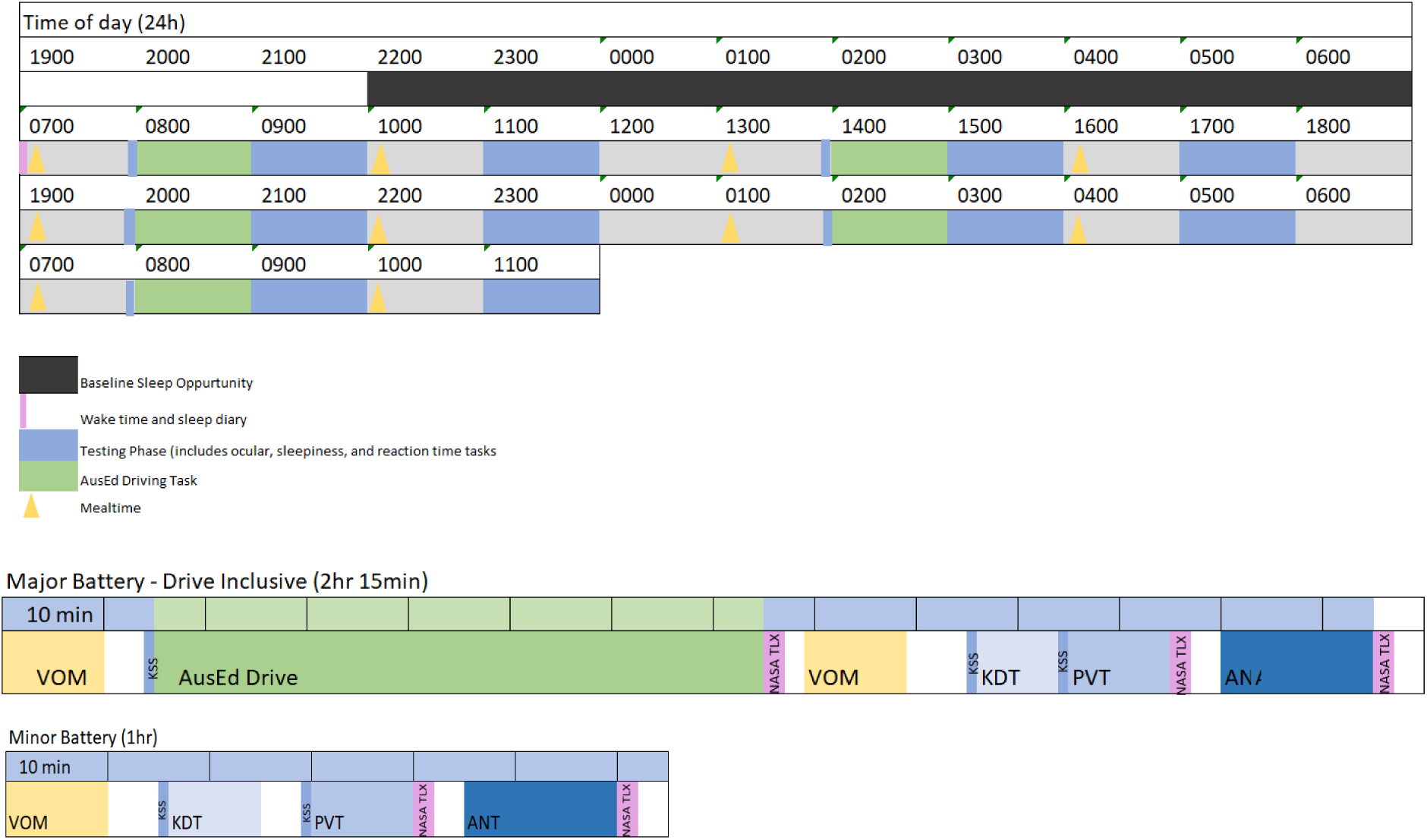
Outline of testing battery timings with example 9-hour bed period during the laboratory stay.

## 3 Statistical analyses

### 3.1 Statement of Prior Statistical Analysis Planning

The analyses were carefully considered and planned before being conducted.

### 3.2 Data Exploration/ Missing Data

The statistical software programs SPSS (version 28.0.1.1 (15) IBM SPSS ©) and R will be used to conduct the statistical analyses. In cases where the data is not normally distributed, data transformations will be performed. In the event that a participant discontinues their participation in the study, or a measurement is missing, an intent to treat analysis will be favoured using all available data. However, individuals without sufficient data will be excluded from the repeated measures tests.

#### 3.2.1 Determining VOM Predictors of Driving Performance

The VOM produced 61 variables relevant for predicting sleepiness (see Supplementary File for variable list). One of the purposes of this presented project was to determine the strongest predictors of sleepiness to enable simplified roadside assessment testing. Variables will be first assessed for correlations between driving performance variables and possible VOM predictors. To rank the importance of each VOM metric in contributing to the model’s predictions, we utilized Shapley values (SHAP) (Lundberg et al., 2020), a state-of-the-art method for interpreting machine learning models. Higher absolute Shapley values indicate greater importance of a VOM metric. Unlike other methods for ranking feature importance, SHAP not only ranks features but also provides insight into the dose-response relationship of a VOM metric’s impact on driving impairment risk. This approach allows us to infer risk thresholds for VOM metrics based on their dose-response relationships. The Gini index will also be compared to a gain metric to assess predictability of variables which is derived from the XGBoost model. XGBoost is widely recognised as a powerful machine learning model and has demonstrated excellent performance across various tasks. Gain is a measure that indicates the importance of predictors. A higher Gain value suggests a greater importance of the predictors. The XGBoost model is recognised as a highly powerful model and has previously demonstrated excellent performance across various research tasks (Asselman et al., 2021). In our research we will use the XGBoost model as the most powerful model. and will run additional models including the Random Forest approach as a fidelity check to increase our confidence in the results from the XGBoost model.

#### 3.2.2 Predicting Driver Sleepiness State

VOM assessments taken immediately before and after each of the five drive simulations will be used, in separate models, to determine the top VOM predictors of sleepiness and driving performance. The VOM test before driving reflects an assessment of fitness to drive before driving begins and the VOM after the drive reflects road-side testing scenario. The daytime drives (drives undertaken at 1, 6 and 13hrs post awakening) will be collated into one model and nighttime drives (drives undertaken at 19 and 25hrs post awakening) will be collated into one model to increase the amount of data available for analysis. The datasets (daytime and nighttime driving sets) will be divided into 80% for model training and 20% for performance evaluation.

Once the most important predictors are determined using SHAP values and XGBoost models, receiver operating curve (ROC) analyses will determine the effectiveness of these variables to predict alertness failure. Alertness failure will be defined through the combination of driving and PVT variables in a twostep cluster analysis to produce a binary outcome for the ROC curves including a vulnerable and a resistant group. The ROC analysis will be performed 100 times and a mean curve and 95% confidence intervals will be extracted to improve our confidence in the predictive ability of the metrics used.

#### 3.2.3 Predicting Trait Sleepiness Future Driving Impairment

To determine whether the VOM can predict future driving impairments, the VOM assessments undertaken prior to the daytime drives will be collated (1hr post waking, 7hrs post waking and 13hrs post waking). The XGBoost models will determine the most important predictors for future driving and alertness performance at 19hrs and 25hrs into wakefulness. It is important to note that the model is trained on the daytime data and then tested on novel data (the nighttime data) to prevent overfitting. The identified predictors will be used to predict future nighttime driving and alertness impairments using ROC analyses. Performance failure for this analysis will be based on data from the alertness testing undertaken at the anticipated circadian nadir (19hrs into wakefulness) at which point performance is theoretically expected to be worst and groups will be defined using a two-step cluster producing a binary outcome variable for the ROC analysis of vulnerable and resistant groups. The ROC analysis will be performed on multiple iterations 100 times and the final accepted ROC curve will be based on the mean of these iterations allowing increased confidence in the outcome. The AUC obtained from the 100 runs of the model will be extracted which will allow us to determine with confidence the predictive power of the top 10 ranked predictors to determine driving impairment.

### 3.3 Secondary Analyses

Further analyses will consider subjective sleepiness, objective sleepiness, and wake electroencephalography using linear mixed modelling techniques or generalised linear mixed effects models in SPSS or R statistical processing software as appropriate. These statistical models will employ an alpha significance level of 0.05 with adjustments for multiple comparisons implemented as appropriate. Relevant effect sizes will be used (partial eta squared or Cohens D) to demonstrate the size of any significant statistical outcomes and increase confidence in *p*-value significance.

## Discussion

This paper presents a new protocol to determine the sensitivity of VOM test metrics to driver state and trait sleepiness and consequent driving simulator impairment during extended wakefulness with the aim of defining the best VOM predictors of driver sleepiness and driving impairment. This is one of the first protocols to directly test a VR device, which comprehensively assesses VOM metrics, to define the best VOM predictors in the context of sleepiness and sleepiness-related driving impairment to address the need for rapid, standardised assessment protocols that can be administered.

VOM measures evaluate a range of gaze parameters arising from vestibular ocular reflex (VOR movements of the eyes to maintain focus on salient features within the visual field (Yorke et al., 2017). Research on the relationship between VOM measures, sleepiness and driving are limited and to date, VOM assessments in the context of driving have been constrained to specialized laboratory environments and focused on individual VOM domain assessments. This limits the feasibility for future deployment and testing of VOM assessment on the road or in the field to identify driver sleepiness. The available experimental evidence suggests there is an increase in VOR gain (the ratio of eye to head angular velocities) during prolonged driving (Kono et al., 2023). Another study found that saccades and anti-saccade were impaired in older patients with poor on-road driving assessment compared with age matched and younger drivers with no driving impairment (Schmitt et al., 2015). Other oculomotor indicators, such as eye blink duration (Filtness et al., 2014) and pupil movement (Malathi et al., 2023), are physiological measures that have also been found useful in assessing driver sleepiness. Despite these observed relationships, no study has previously utilised a mature VR technology device, which would be readily deployable in the field, to comprehensively assess VOM metrics in relation driver sleepiness and driving impairment. The current study protocol addresses this important gap.

The study was warranted as it seeks to test the potential of a practical approach to detecting driver alertness failure that can, with future studies, be replicated and potentially implemented in real-world roadside and transportation workplace contexts to reduce the risk of road accidents. In addition, detection of trait alertness may support prognostic evaluation of vulnerability to sleep loss and driving impairment, which could be deployed in clinical or occupational fitness to drive screening. Having better insights into sleepiness state would allow provide licence drivers as well as professional drivers and employees to make more informed decisions regarding sleepiness counter measures including work/rest scheduling (Cori et al., 2021), caffeine and strategic naps(Horne & Reyner, 1996; Wörle et al., 2021) and lighting interventions (Sletten et al., 2017). The current study seeks to make an important contribution to the field of road safety testing and provide valuable insights for alertness monitoring and driving risk prediction.

A major strength of this study was the use of gold standard laboratory testing methods where environments are controlled, and the influence of extraneous variables could be monitored and controlled. This was crucial in this early stage of determining efficacy of this approach. This project will provide the first steps into understanding the usefulness of the VR VOM device and its predictive power and if successful will subsequently inform infield on-road testing in uncontrolled environments towards real world validation. There are potential limitations that would need to be considered. Although our sample size of 51 participants may appear small, it significantly exceeds sample sizes achieved by similar extended wakefulness studies (Bratzke et al., 2009; Bratzke et al., 2012). Given our repeated measures design with 5 driving assessments throughout the extended wakefulness, as well as established effect sizes for the primary driving simulator test(Desai et al., 2007; Vakulin et al., 2007; Vakulin et al., 2009; Vakulin et al., 2014; Vakulin et al., 2016) ensures that we have sufficient power when developing VOM prediction models. However, any findings would require independent validation. Another possible limitation to deployment of the VOM test for sleepiness is the test duration and test parameters. This will be informed by this study to allow the development of a more focused and targeted test including only the VOM metrics that are useful for sleepiness and driving impairment detection. Furthermore, there are ecological limitation of the current device which requires refinement for practical field use, particularly regarding device weight, connectivity and portability which would need to be resolved for in-field applications. Once the core protocols and prediction algorithms developed are thoroughly tested and validated, subsequent engineering iterations can address these test parameter and hardware implementation challenges.

In conclusion, by identifying an objective measure of sleepiness-related impairment, this work contributes to addressing a critical gap in road safety where a road-side sleepiness test and/or fitness to drive assessments are currently lacking. This is particularly relevant for populations who face unavoidable sleepiness risks, such as emergency personnel and transportation shift workers and new parents, where traditional countermeasures such as flexible scheduling may be impractical or impossible. Similar to how alcohol testing has incentivised responsible drinking behaviour, objective sleepiness detection could encourage better fatigue management behaviours for private drivers, provide decision support of employees and employers in transportation shift work industry, and for clinicians assessing fitness to drive in drivers with sleep disorders.

## Data Availability

This manuscripts descibes the protocol for an extended wakefulness study and hence no data is presented

## Acknowledgements

The authors acknowledge the Commonwealth Department of Infrastructure, Transport, Regional Development and Communications, Office of Road Safety grant Road Safety Innovation Fund –96 (RSIF2-96) for their financial funding of this research study.

In addition, the researchers would like to thank all participants who have been involved in this research allowing us to improve the safety of our roads. They further acknowledge the research assistants and students who continue to work to make this project a success.

The investigators would also like to thank NeuroFlex the industry partner on this Grant for supplying the virtual reality devices which allows for the assessment of the primary variable of interest, vestibular ocular motor reflexes.

## Author Contributions

AG, CD, NL, KN, KB, DPN, AR, RA, LL, PC, JC, MEH, CA, DB, AM, NS and AV developed the study concepts and aims. All authors provided important insight on data interpretation and drafting of the manuscript. All authors approve of the final version of the manuscript.

## Declarations of interest

AG, KN, DPC, LL, DBB, AM, NS, LL have no conflicts of interests to declare.

AV research supported by the National Health and Medical Research Council project grants and fellowship. He has received competitive funding and equipment from ResMed, Philips Respironics Withings and Sleeptite for research unrelated to this project. He has received Neuroflex devices directly to support this project with Neuroflex being a formal industry partner on this project. Unrelated to the current project AV reported Decision Support Software System for Sleep Disorder Identification Decision support algorithms for online sleep disorder screening survey. Being a Leadership Academic Chair – Sleep Health Foundation Consumer Reference Panel, Chair - Steering Committee, Australian Sleep and Alertness Consortium (ASAC), Board member – Sleep Health Foundation, Board member – Australasian Sleep Association, Various institutional and professional leadership, unsalaried and unrelated roles to this manuscript.

DS is the industry partner representative on this project but was not involved in the current study protocol design, data collection or analysis. The project was investigator led and initiated, with no involvement from Neuroflex in the study design, data collection or data analysis.

HS reports receiving Support for manuscript Commonwealth Department of Infrastructure, Transport, Regional Development and Communications Road Safety grant Road Safety Innovation Fund – 96 (RSIF2-96) NeuroFlex Supplied devices for testing. Received grants and equipment from American Academy of Sleep Medicine Foundation Research, Re-Time Pty Ltd In-kind contribution, Compumedics Ltd In-kind contribution and withings Sleep Analyser devices for research purposes unrelated to the project. Flinders University also supplied research funding for travel unrelated to this work.

KB reports drawing salary from the RSIF grant which funded this project. ACR reports research income from Sydney Trains, Compumedics, and the Sleep Health Foundation for research related to sleep and sleep disorders.

PC reports grants unrelated to this study from National Health and Medical Research Council, Defence Science and Technology Group, Flinders University, Compumedics Ltd, Invicta Medical, Flinders Foundation, Medical Research Future Fund, Garnett Passe and Rodney Williams Memorial Foundation and Minister for Innovation and Skills, South Australia/Science, Technology and Commercialisation, and in-kind support of unrelated trials from REDARC and Re-Timer Pty Ltd.

RB reports grants from National Health and Medical Research Council, ResMed Foundation, The Hospital Research Foundation, Philips Respironics, Australian Government, Defence Science and Technology Group, Flinders Foundation all unrelated to the current trial. A Sleep Health Foundation unpaid board member and received In-kind Equipment from Philips Respironics unrelated to this project.

JC and MH reports grants unrelated to this project from Australian Automobile Association Jack Brockhoff Foundation and Department of Transport and Planning National Transport Commission. Also, payment for expert testimony Transport Safety Victoria from Seeing Machines and Optalert. MH also reported being an Honorary board member at the Institute for Breathing and Sleep.

AR reported the following all unrelated to the current project; grants from Lifetime Support Authority (Government of SA) Research, Sydney Trains Research, and Flinders Foundation Research Funding to institution. And received Consulting fees Compumedics Ltd Consulting. Speaker presentation funds from Teva Pharmaceuticals, Sleep Health Foundation, Research Review, and Governing Council.

NL reported the following all unrelated to the current project; grants from National Health and Medical Research Council of Australia, Philips Respironics Research, The Hospital Research Foundation Research, ResMed Foundation Research. Equipment from ResMed, Philips Provision.

CD reported the following all unrelated to the current project; grants from development Student university Grant awarded 2020 for rural sleep health seminars, Funded a student led outreach program HDR/ECR representation for executive meetings for the FHMRI:Sleep Health institute Volunteer role, and received The Hospital Research Foundation – THRF PhD scholarship awarded in 2019. CD further reports drawing salary from the RSIF grant which funded this project.

C.A. has received a research award/prize from Sanofi-Aventis; contract research support from Department for Transport Victoria (formerly VicRoads), Transport Accident Commission, Rio Tinto Coal Australia, National Transport Commission, Tontine/Pacific Brands, and the Australian Automobile Association; industry funding through ARC Linkage scheme with Seeing Machines and Cogstate Ltd; and lecturing fees from Brown Medical School/Rhode Island Hospital, Ausmed, Healthmed and TEVA Pharma Australia; and reimbursements for conference travel expenses from

Philips Healthcare. In addition, she has served as a consultant to the Rail, Bus and Tram Union, the Transport Accident Commission (TAC), the National Transportation Committee (NTC), VicRoads/Department of Transport Victoria, and Melius Consulting. She has also served as an expert witness and/or consultant in relation to fatigue and drowsy driving and was a Theme Leader in the Cooperative Research Centre for Alertness, Safety and Productivity. These conflicts are unrelated to the data presented in this manuscript.

