## Supplementary Tables 1 to 4 for "Evaluating the effectiveness of vestibular and ocular motor function assessments in detecting driver sleepiness: A Protocol Paper"

**SUPPLEMENTARY FILE**

### Determining the effectiveness of vestibular and ocular motor function assessments to identify excessively sleepy drivers: Protocol Paper

Alisha Guyett[1#], Claire Dunbar[1#], Nicole Lovato[1], Katrina Nguyen[1], Kelsey Bickley[1], Duc Phuc Nguyen[1], Amy Reynolds[1], Maslin Hughes[1], Hannah Scott[1], Robert Adams[1], Leon Lack[1], Peter Catcheside[1], Lucia Pinilla[1], Jennifer Cori[2,3], Mark E. Howard[2,3,4], Clare Anderson[3,5], David Stevens[1,6], Darah-Bree Bensen-Boakes[1], Ashley Montero[1], Nikki Stuart[1], Andrew Vakulin[1]

### co-first authors

1. Flinders Health and Medical Research Institute: Sleep Health, Flinders University, Adelaide, Australia.
2. Institute for Breathing and Sleep, Austin Health, Heidelberg, Victoria, Australia.
3. School of Psychological Sciences and Turner Institute for Brain and Mental Health, Monash University, Clayton, Victoria, Australia.
4. Department of Medicine, The University of Melbourne, Parkville, Victoria, Australia
5. Centre for Human Brain Health, School of Psychology, University of Birmingham, Edgbaston, UK
6. Digital Health Organisation (subsidiary of Saccade Analytics, Inc.)

**Correspondence:**
Alisha Guyett
Flinders University, Mark Oliphant Building, Flinders Health and Medical Research Institute: Sleep Health
5 Laffer Drive, Bedford Park, South Australia 5042


Number of words: 1281

Number of Tables: 2
Number of Figures: 1

#### Supplementary 1:

**Vestibular Ocular Motor protocol descriptions**

The instructions for all tests appear inside the goggles for the participants, as well as on the screen for the tester. This minimised intraindividual differences in test delivery. The stimulus for all tests appears at a 1m simulated distance. Furthermore, the stimuli presentation for each test is randomised to minimise learning impact. Prior to the test, a 15s calibration occurs which allows the infrared sensors to accurately detect the iris and pupil.

1. **Smooth pursuit (free head)**

During visual tracking, the eyes and head follow a slowly moving central target in both horizontal (H) and vertical (V) directions. This coordinated movement occurs within a wide field of view, approximately ± 80 degrees horizontally and vertically. The target remains at a fixed distance from the head, eliminating the need for vergence adjustments. To prevent predictability, the target's trajectory incorporates random elements. Periodic corrective jerks are made by the subject to keep the target in focus and minimize deviations. In healthy individuals, eye movements are predominantly coordinated, with minimal involvement of vergence. However, deviations exceeding 4 degrees, whether vergence or conjugate, may impair clear vision as they exceed the foveal range.

2. **Smooth pursuit (fixed head)**

During the visual tracking assessment, whether conducted with a fixed head the patient is tasked with continuously maintaining a moving target at a distance of 1 meter. When the test is performed with the head free, coordinated movements of the head and eyes are employed to keep the target centred within the visual field. This test effectively integrates visual and vestibular systems, along with neck proprioception, offering a comprehensive evaluation of functional movements. With a stationary head, the target typically remains within a ± 30-degree radius. Alternatively, pursuit with the eyes alone isolates oculomotor movements, making it suitable for patients with limited cervical mobility. Vergence, the difference in gaze angle between the eyes, is typically minimal or absent in healthy individuals, indicating convergence or divergence. The random trajectory of the target prevents predictability and subsequent biases. Ideally, the average tracking error should not exceed 3.5 degrees. Significant variations in vergence or tracking errors exceeding 3.5 degrees, along with excessive corrective jerks, suggest weaknesses in the visual tracking system.

3. **Active visual VOR (horizontal)**

This test assesses the active ocular vestibulo-visual reflex in either horizontal or vertical direction. The protocol involves integrating visual and vestibular cues with neck proprioception, mimicking natural conditions encountered in daily activities. In contrast to traditional response vector orientation (RVO) tests where head rotation is passive and conducted in darkness, this test incorporates more sensory input to enhance performance. By combining these sensory inputs, individuals can better perceive visual objects with clarity. The test evaluates three main aspects: gain, average vergence, and glide speed between the retina and the target positioned 1 meter away. In healthy individuals, there is minimal vergence and a glide speed of less than 4 deg/s, associated with gains of 80% or more. In cases of deficient RVO, where the eyes fail to compensate for head movements effectively, vision can become blurred and vertigo may occur. If the slip speed of the image on the retina exceeds 4 deg/s, individuals may struggle to distinguish letters for reading. Moreover, varying vergence in relation to head speed can indicate vestibular weakness. For clear vision, the error between target position and gaze direction during conjugate movements should not exceed 4 degrees, while an average vergence exceeding 4 degrees hampers clear vision by surpassing foveal accuracy.

**4. Active visual VOR (vertical)**

This test evaluates the Rotatory Visual Vestibulo-Ocular Adaptation in either the horizontal or vertical direction. The protocol involves integrating visual and vestibular cues with neck proprioception, simulating natural conditions encountered in everyday activities. In contrast to traditional RVO tests where head rotation is passive and conducted in darkness, this test incorporates more sensory input to enhance performance. By combining these sensory inputs, individuals can better perceive visual objects with clarity. The test assesses three main aspects: gain, average vergence, and glide speed between the retina and the target positioned 1 meter away. In healthy individuals, there is minimal vergence, and the glide speed is 4 deg/s less, associated with gains of 80% or more. In cases of deficient RVO, where the eyes fail to compensate for head movements effectively, vision can become blurred and vertigo may occur. If the slip speed of the image on the retina exceeds 4 deg/s, individuals may struggle to distinguish letters for reading. Moreover, vestibular weakness is confirmed if vergence varies in association with head speed. During conjugated movements, the error between the position of the target and the direction of gaze should not exceed 4 degrees for clear vision. Additionally, an average vergence above 4 degrees impedes a clear view of the target by surpassing foveal precision.

**5. Saccades**

This assessment evaluates eye performance during random saccades, which are rapid jumps to new targets, while the head remains stationary. Random targets are presented within the subject's visual field (< 30 degrees) at a consistent virtual distance. The analysis focuses on three key aspects of these jumps: reaction time, precision, and vergence, aiming to identify deficits in specific regions of the visual field. The standard reaction time for these jumps is around 150ms. When eye movements are well-coordinated, vergence measures are minimal, ideally remaining below 4 degrees to ensure clear visibility of the target.

**6. Anti-Saccades**

In this test, a brief presentation of a red target occurs horizontally for the subject. However, the task demands that the subject looks not at this target but instead at an imaginary point equidistant from the centre but in the opposite direction (e.g., if the target appears 10 degrees to the right, the subject must look 10 degrees to the left). This adds a cognitive aspect along with rotations of the visual plane before executing the movement. It is often employed to assess visual cortex function in patients with head trauma. Deficits typically manifest as increased reaction times and deviations compared to regular jerks performed by the same subject. Deviations beyond 4 degrees indicate a lack of accuracy required for clear image perception.

**7. Optokinetic Nystagmus**

Optokinetic Nystagmus occurs when the subject's eyes track one of several targets that are scrolling together. To elicit this response, a field of dots moves harmonically, initially horizontally and then vertically. OKN nystagmus comprises slow phases of eye movement, which track the visual field, interspersed with rapid jerks that enable the subject, upon fixating on a new object, to redirect their gaze back toward the center. The results of this test are assessed similarly to those of Rotatory Visual Vestibulo-Ocular Adaptation and typically yield comparable outcomes in healthy individuals, provided the peripheral retina's vision remains intact.

**8. Spontaneous Nystagmus**

Spontaneous nystagmus, an involuntary eye movement occurring without external visual, vestibular, or cognitive stimuli, serves as an indicator of imbalance within the gaze orientation system. This imbalance may result from head trauma, vestibular dysfunction, neurodegenerative disorders, or genetic factors. During the test, a target briefly appears in the subject's field of vision before vanishing, prompting the subject to maintain fixation at the target's last location for a few seconds in darkness. Multiple locations in the visual field are assessed. Key measures include the frequency of involuntary jerking, eye drift, and tremor (provided for reference). The location with minimal measurements denotes the preferred 'resting' position, which may not align directly in front of the head. A frequency exceeding 2 saccades per second can induce blurred vision, as it lacks the approximately 350 milliseconds required for a physiological slow-phase response. Eye drift occurs when the gaze deviates from the preferred zero point; if the drift rate exceeds 4 degrees per second, vision becomes blurred.

**Supplementary 2:**

Vestibular Ocular Motor measure variables calculated from assessments and included in analysis for prediction models.

| | **Metric** | **Explanation** |
|---|---|---|
| **Smooth pursuit (*'head free'*, or head and eyes together)** | Mean (ocular) vergence (°) | Measure of strabismus (how 'parallel' the eyes are), specifically estotropia (eyes turned inwards, or 'cross-eyed') or exotropia (eyes turned outwards). |
| | Ocular vergence Standard Deviation (°) | Measure how variability of strabismus, as in the magnitude the eyes go between esotropia and exotropia to maintain focus on the stimuli. *As the stimuli is set a 1m distance, the standard deviation should be minimal.* |
| | Mean Error (°) | How far away from the stimuli the gaze was focussed. |
| | Number of Saccades | How many 'corrective saccades' occurred to keep track of the stimuli. *During smooth pursuit, saccades should be minimal.* |
| | Head Contribution (Mean) (%) (also calculated indicividually for horiztonal and vertical planes) | The proportion of contribution made by the head. *100% contribution indicates the head and eyes co-ordinated perfectly. <100% indicates eye predominance. >100% indicates head predominance.* |
| **Active visual VOR (both horizontal and vertical)** | Mean (ocular) vergence (°) | Measure of strabismus (how 'parallel' the eyes are), specifically estotropia (eyes turned inwards, or 'cross-eyed') or exotropia (eyes turned outwards). |
| | Ocular vergence Standard Deviation (°) | Measure how variability of strabismus, as in the magnitude the eyes go between esotropia and exotropia to maintain focus on the stimuli. *As the stimuli is set a 1m distance, the standard deviation should be minimal.* |
| | Gain (%) | The ratio between head and eye movement. A gain of 100% indicates for every 10° the eyes move, the |

| | | |
|---|---|---|
| | | head compensates by 10°. <100% indicates eye predominance. >100% indicates head predominance.<br>NB gain is calculated for 25°.sec$^{-1}$, 50°.sec$^{-1}$, and 75°.sec$^{-1}$ |
| **Smooth pursuit (*'head fixed'*, or eyes only)** | Mean (ocular) vergence (°) | Measure of strabismus (how 'parallel' the eyes are), specifically estotropia (eyes turned inwards, or 'cross-eyed') or exotropia (eyes turned outwards). |
| | Ocular vergence Standard Deviation (°) | Measure how variability of strabismus, as in the magnitude the eyes go between esotropia and exotropia to maintain focus on the stimuli. *As the stimuli is set a 1m distance, the standard deviation should be minimal.* |
| | Mean Error (°) | How far away from the stimuli the gaze was focussed. |
| | Number of Saccades | How many 'corrective saccades' occurred to keep track of the stimuli. *During smooth pursuit, saccades should be minimal.* |
| **Prosaccades** | Mean (ocular) vergence (°) | Measure of strabismus (how 'parallel' the eyes are), specifically estotropia (eyes turned inwards, or 'cross-eyed') or exotropia (eyes turned outwards). |
| | Ocular vergence Standard Deviation (°) | Measure how variability of strabismus, as in the magnitude the eyes go between esotropia and exotropia to maintain focus on the stimuli. *As the stimuli is set a 1m distance, the standard deviation should be minimal.* |
| | Mean Error (°) | How far away from the stimuli the gaze was focussed. |
| | Mean latency (ms) | Reaction time to looking at the stimuli after it has moved. |
| **Antisaccades** | Mean (ocular) vergence (°) | Measure of strabismus (how 'parallel' the eyes are), specifically estotropia (eyes turned inwards, or 'cross-eyed') or exotropia (eyes turned outwards). |
| | Ocular vergence | Measure how variability of strabismus, as in the |

| | | |
|---|---|---|
| | Standard Deviation (°) | magnitude the eyes go between esotropia and exotropia to maintain focus on the stimuli. *As the stimuli is set a 1m distance, the standard deviation should be minimal.* |
| | Mean Error (°) | How far away from the stimuli the gaze was focussed. |
| | Mean latency (ms) | Reaction time to looking at the stimuli after it has moved. |
| | Directional accuracy (%) | How many times the person looked *away* from the stimuli. 100% means the person looked away every time. |
| **Optokinetic nystagmus** | Mean (ocular) vergence (°) | Measure of strabismus (how 'parallel' the eyes are), specifically estotropia (eyes turned inwards, or 'cross-eyed') or exotropia (eyes turned outwards). |
| | Ocular vergence Standard Deviation (°) | Measure how variability of strabismus, as in the magnitude the eyes go between esotropia and exotropia to maintain focus on the stimuli. *As the stimuli is set a 1m distance, the standard deviation should be minimal.* |
| | Gain % (all directions – left, right, up, and down) | The ratio between eye movement velocity, and the stimuli velocity. 100% means for eyes matched the velocity of the stimuli. Under 100% mean the eyes were slower than the stimuli. Over 100% meant the eyes were faster. Calculated for 25°.sec$^{-1}$ and 50°.sec$^{-1}$. |
| **Spontaneous nystagmus (often referred to as *'gaze evoked nystagmus'*).** | Mean tremor frequency (saccades.sec$^{-1}$) | Number of saccades.sec$^{-1}$ whilst gazing. >0.5 saccades.sec$^{-1}$ indicates nystagmus. Measured individually for central gaze, and left/right/up/down peripheral gaze. |
| | Mean tremor velocity (°.sec$^{-1}$) | Average velocity of saccades during gazing. |
| | Mean drift (°.sec$^{-1}$) | The ratio between eye movement velocity, and the stimuli velocity. 100% means for eyes matched the velocity of the stimuli. Under 100% mean the eyes |

| | | |
|---|---|---|
| | | were slower than the stimuli. Over 100% meant the eyes were faster. Calculated for 25°.sec$^{-1}$ and 50°.sec$^{-1}$. |

**Table 1 Vestibular Ocular Motor measure variables calculated from assessments and included in analysis for prediction models.**

#### Supplementary 3:

Key of test battery timings and tasks undertaken during the major and minor batteries.

Key of test battery timings and tasks undertaken during the major and minor batteries.

| Performance task | Time from wake (hh.mm) |
|---|---|
| **Major Battery** | |
| VOMs Battery (10 minutes)<br>Aus-Ed pre drive KSS | +00.30<br>+00.43 |
| Aus-Ed Drive (60 minutes) | +00.45 |
| Aus-Ed Post-Drive NASA-Task Loading Index | +01.45 |
| VOMs Battery (10 minutes) | +01.50 |
| KDT (w/KSS pre and post) | +02.05 |
| Psychomotor Vigilance Test (PVT) (10 minutes) | +02.15 |
| Post-PTV NASA-Task Loading Index | +02.25 |
| Attention Networking Task (ANT) (10 minutes) | +02.30 |
| Post-ANT NASA-Task Loading Index | +02.45 |
| Battery 1 part 1 completed, participant free time | |
| Meal | +03.00 |
| **Minor Battery** | |
| VOMs Battery (10 minutes) | +04.00 |
| KDT (w/KSS pre and post) | +04.15 |
| PVT (10 minutes) | +04.30 |
| Post-PVT NASA-Task Loading Index | +04.40 |
| ANT (10 minutes) | +04.45 |
| Post-ANT NASA-Task Loading Index | +05.00 |
| Battery 1 part 2 completed, participant free time | |

**Table 2. Overview of one 6 hour period including Major and Minor battery components and all tests and assessments undertaken during this time. This protocol was repeated five times throughout the extended wakeful*ness.***

#### Supplementary file 4:

| Assessments | Study Period | | | |
|---|---|---|---|---|
| | Screening | Pre-Laboratory Visit | Laboratory Protocol | |
| | | | Adaptation Night | Extended wake period |
| Screening Assessments | | | | |
| General Health, Medical and Lifestyle Screening Questionnaire | X | | | |
| Inclusion/Exclusion Criteria | X | | | |
| Enrolment | X | | | |
| Informed Consent | X | | | |
| Pre-Laboratory Monitoring | | | | |
| Withings | | X | | |
| Gene Actiwatch | | X | | |
| Sleep Diaries | | X | | |
| Sleep Monitoring | | | | |
| Sleep Diaries | | X | X | |
| Gene ActiWatch | | X | X | X |
| Polysomnography | | | X | X |
| Biological Sample Collection | | | | |
| Core Body Temperature | | | X | X |
| Health Assessment | | | | |
| Height, Weight, Waist, Hips | | | X | |
| Blood Pressure | | | X | |
| Wake Heart Rate | | | X | |
| Neurocognitive Tasks | | | | |
| Karolinska Drowsiness Test | | | | X |
| Psychomotor Vigilance Test – 10-minute | | | | X |
| Vestibular Ocular Montor Assessment | | | | X |
| Aus-Ed Drive | | | | X |
| Attention Network Test | | | | X |
| Questionnaires | | | | |
| Epworth Sleepiness Scale | X | | | |

| | | | | |
|---|---|---|---|---|
| Insomnia Severity Index | X | | | |
| Obstructive Sleep Apnoea 50 | X | | | |
| Good Sleeper Index | X | | | |
| Morning-Eveningness Questionnaire | X | | | |
| Pittsburgh sleep Quality Index | X | | | |
| Sheehan Disability Scale | X | | | |
| Depression, Anxiety and Stress Scale | X | | | |
| Karolinska Sleepiness Scale | | | | X |
| NASA Task Loading Index | | | | X |

**Table 2. All Study Assessments and Timing of Administration.**
